# Stability of Microbiome-Derived Fatty Acids in Self-Collected Samples: A Comparative Evaluation of Stool and Blood Matrices

**DOI:** 10.64898/2026.03.05.26347712

**Authors:** Matteo Domenico Marsiglia, Michele Vittorio dei Cas, Silvia Bianchi, Elisa Borghi

## Abstract

**Background:** Short-chain fatty acids (SCFAs) are widely used as functional readouts of gut microbial activity *in vivo*. The growing adoption of decentralised study designs and self-collection protocols has amplified the need for reliable room-temperature storage and shipment strategies. However, SCFAs volatility and the persistence of post-collection microbial metabolism raise concerns regarding pre-analytical stability and the interpretability of measured concentrations.

**Methods:** We assessed the temporal stability of fatty acids (FAs) across intestinal and systemic matrices under room-temperature storage. Untreated stool was compared with two nucleic acid stabilisation devices (eNAT and OMNIgene-GUT), while whole blood, plasma and dried blood spots (DBS) were evaluated as minimally invasive systemic sampling strategies. Profiles were quantified using complementary GC-MS and LC-MS/MS workflows.

**Results:** Untreated stool showed fermentation-driven increases in major SCFAs, whereas immediate freezing preserved baseline profiles. eNAT maintained faecal FA stability for up to 21 days, while OMNIgene-GUT exhibited baseline and time-dependent alterations. In systemic matrices, plasma and whole blood showed upward drift, whereas DBS declined initially before stabilising after approximately 14 days.

**Conclusions:** FA measurements are highly matrix- and device-dependent. Our findings provide practical guidance for the selection of sampling strategies in microbiome-associated FA studies and emphasise the need for controlled pre-analytical conditions in decentralised microbiome studies.

## INTRODUCTION

Short-chain fatty acids (SCFAs), primarily acetate, propionate and butyrate, constitute a major functional output of the gut microbiota and are among the most widely used proxies of microbial metabolic activity *in vivo.* Produced through the anaerobic fermentation of dietary substrates by diverse bacterial taxa, SCFA concentrations reflect not only microbial community composition but also substrate availability and metabolic flux, thereby linking taxonomic structure to realised function. For this reason, SCFAs are frequently used as functional endpoints in microbiome research, including longitudinal studies, dietary interventions and cross-cohort comparisons, where they are often interpreted as indicators of microbial activity rather than mere downstream host metabolites.^1–3^

Unlike sequencing based readouts, which capture genetic potential, SCFA profiles represent an integrated phenotype emerging from complex host-microbiota interactions. Large scale microbiome-metabolome studies have demonstrated that microbial metabolites, including SCFAs, do not map linearly onto individual taxa but instead arise from community level interactions and shared metabolic pathways, reinforcing their role as functional bridges between microbial composition and biological outcome.^4–6^ Consequently, SCFA measurements are increasingly used to support causal or mechanistic biological interpretations of microbiome activity.

To date, no universally accepted standardized collection protocols have been established to reliably preserve SCFA content in biological matrices, which may affect the accuracy and comparability of microbiota-derived metabolic data. SCFAs are among the most analytically fragile microbiome-associated metabolites due to their volatility, chemical reactivity and susceptibility to ongoing microbial or enzymatic activity. Accumulating evidence indicates that pre-analytical variables, including sample matrix, collection procedures, storage temperature and duration, can induce substantial compound-specific drift in SCFA abundance often exceeding underlying biological variability.^7–9^ Recent analytical studies further highlight that even short delays between sampling and processing can significantly alter SCFA profiles, particularly in stool and low-concentration blood-derived matrices.^10–13^

These challenges are amplified in the context of large-scale and decentralised microbiome studies, where immediate processing is rarely feasible. Conventional stool collection is logistically demanding and prone to temperature- and time-dependent metabolic changes, while circulating SCFAs occur at low concentrations that are highly sensitive to adsorption, evaporation and oxidative loss. Although advances in Gas Chromatography–Mass Spectrometry (GC–MS) and Liquid Chromatography–Mass Spectrometry (LC–MS) methodologies have markedly improved analytical sensitivity and specificity, improved detection alone cannot recover biological information lost through pre-analytical degradation.^14–17^ As a result, differences in SCFA abundance across studies may reflect methodological artefacts rather than true biological variation.

To address these limitations, alternative sampling strategies such as stool collection devices with preservatives and dried blood spots (DBS) have gained increasing attention due to their logistical advantages and suitability for remote sampling.^18–22^ Nevertheless, the extent to which these approaches preserve SCFA profiles over time, and whether they introduce systematic, matrix-dependent bias, remains insufficiently characterised. In the absence of validation, the widespread use of SCFA profiling as a functional microbiome readout risk conflating technical variability with biological signal.

In this study, we systematically evaluate the temporal stability of SCFAs and medium-chain fatty acids (MCFAs) across intestinal and systemic matrices under extended room-temperature storage. Specifically, we sought to validate non-invasive self-collection approaches applicable to multicentre and longitudinal study designs, thereby reducing the need for unnecessary in-person clinic visits among participants. We compare untreated stool with two widely used nucleic acid stabilisation devices (i.e. eNAT and OMNIgene-GUT) and assess whole blood, plasma and DBS to determine whether minimally invasive sampling strategies can yield reproducible metabolite profiles. By quantifying matrix- and device-specific drift over time, our work aims to define practical sampling conditions that preserve biologically meaningful SCFA signals and strengthen the interpretability and reproducibility of microbiome–metabolite studies.

## METHODS

### Study design

The study was structured into two complementary experimental arms examining the stability of short-and medium-chain fatty acids under room-temperature (RT) storage across faecal and systemic matrices (Fig. 1).

**Figure 1.**
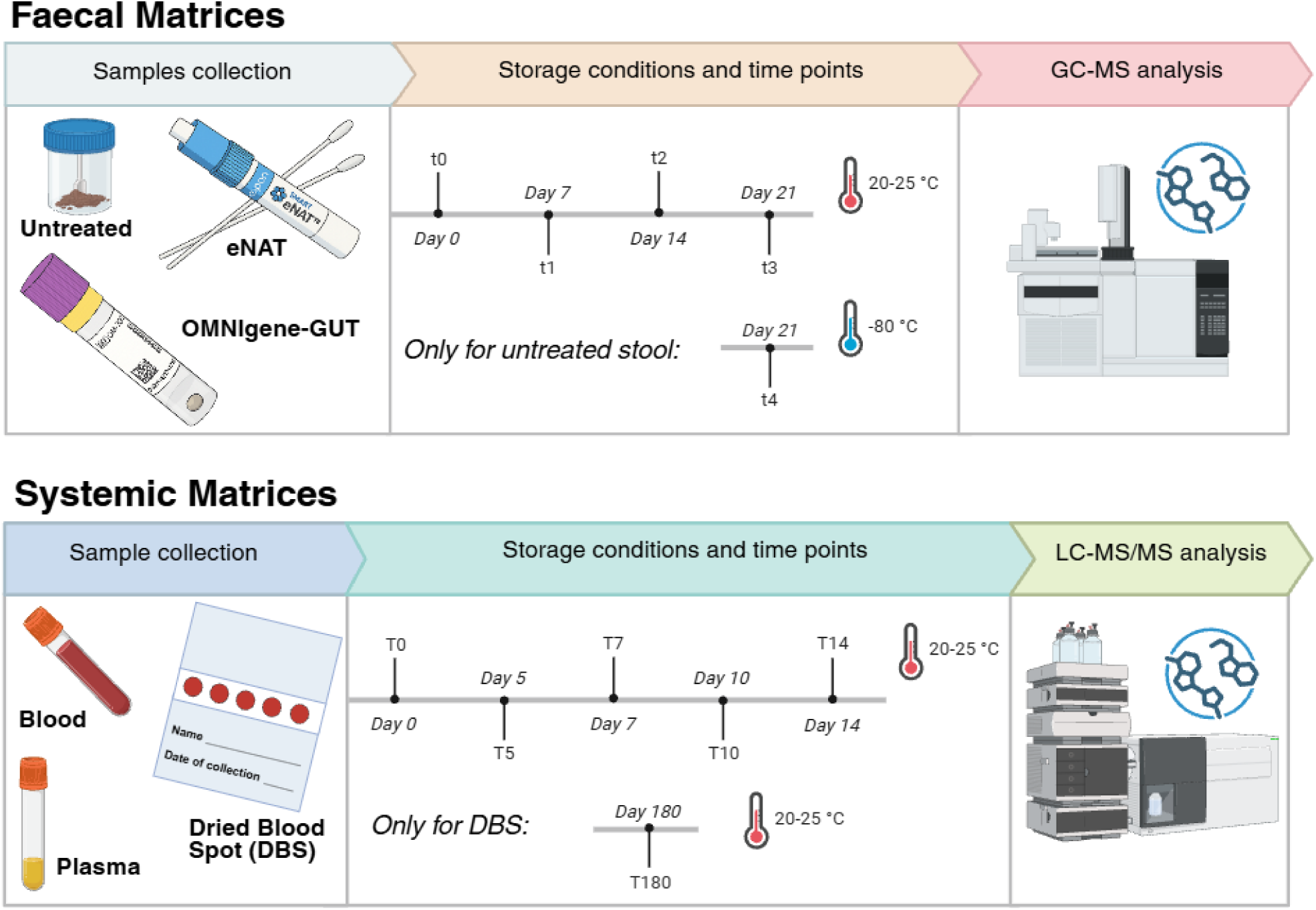
Overview of the study design. Schematic representation of sample collection, storage conditions and analytical workflows used to assess fatty acid stability across faecal and systemic matrices. Created in BioRender. Marsiglia, M. D. (2026) https://BioRender.com/9yzmgpx.

In the faecal arm, stool material was collected and processed using three conditions: untreated stool, eNAT® (Copan), and OMNIgene®•GUT OM-200 (DNA Genotek). Each sample was analysed at baseline (t0) and again after 7 days (t1), 14 days (t2), and 21 days (t3) of storage at RT. For the untreated condition, an additional aliquot collected at t0 was immediately frozen at −80 °C and re-analysed at the end of the study (t4) to serve as a frozen-reference control.

The systemic arm included three blood-derived matrices, whole blood, dried blood spots (DBS; Euroimmun), and plasma, each monitored under the same RT conditions. Samples were analysed at baseline (T0) and after 5 days (T5), 7 days (T7), 10 days (T10), and 14 days (T14) of RT storage. To evaluate long-term preservation performance, the DBS matrix included an extended timepoint at 180 days (T180).

This study was performed in line with the principles of the Declaration of Helsinki. Written informed consent was obtained from each participant following protocols approved by Comitato Etico Territoriale Lombardia 1 (CET 129-2024).

### Fatty acids extraction protocols Stool samples

The extraction of fatty acids (FAs) from frozen stool samples was conducted as follows: faecal samples (200 mg) were homogenized in 1 mL of water. Subsequently, 700 µL of water, 200 µL of pure orthophosphoric acid (85%), and 100 µL of internal standard (2-ethylbutyric acid, 3.3 mM) were added to 300 µL of the homogenate. The mixture was extracted with 500 µL of a diethyl ether/heptane solution (1:1) and gently shaken for 15 minutes. After centrifugation for 5 minutes, the organic phase was recovered for the GC-MS analysis.^23^

### eNAT and OMNIgene-GUT stool samples

Stool samples were weighted and added to eNAT and OMNIgene-GUT devices. A 200 µL of the stool-stabilizing buffer mixture were combined with 800 µL of water, 200 µL of pure orthophosphoric acid (85%), and 100 µL of internal standard. The extraction was performed using the same diethyl ether/heptane protocol as described above.

### Plasma, whole blood and dried blood spots samples

For each subject, venous whole blood was collected into EDTA tubes and immediately processed into three matched matrices: whole blood, DBS and plasma. An aliquot of 9 µL of whole blood was set aside for direct analysis. A second aliquot was spotted onto DBS filter paper and allowed to dry at RT. After drying, circular punches of the DBS cards were obtained using a manual puncher of appropriate diameter (6 mm) to correspond to approximately 9 µL of blood per disc.^24,25^ The remaining blood was centrifuged, the resulting plasma were collected and 9 µL were tested.

Fatty acids were extracted using a protein-precipitation protocol common to all three matrices. Each sample (9 µL of plasma, 9 µL of whole blood or one DBS punch) was mixed with 200 µL of ice-cold isopropanol and incubated for 2 h at 4 °C under gentle shaking. Samples were then centrifuged at 21000 xg for 10 min at 4 °C, and 100 µL of the clear supernatant were transferred to a new glass vial for derivatisation. 100 µL of supernatant were combined with 50 µL of 1-Ethyl-3-(3-dimethylaminopropyl)carbodiimide (EDC) solution (10 mg/mL in 70% methanol), 50 µL of 3-nitrophenylhydrazine (3-NPH; 10 mg/mL in 70% methanol), 50 µL of pyridine (7% v/v in 70% methanol) and 10 µL of an internal standard mixture containing (2-isobutoxyacetic acid, 2.5 μg/mL and undecanoic acid,10 µg/mL). The reaction mixture was incubated for 1 h at 40 °C in a thermoblock. The derivatization was stopped by the addition of 4 µL of 2.5% formic acid, vortexed briefly and transferred to LC–MS/MS vials for the analysis.

### Fatty acids quantification GC-MS analysis

The quantification of faecal acetic, propionic, isobutyric, butyric, isovaleric, valeric, caproic, caprylic, capric acids were assessed by gas chromatography-mass spectrometry (GC-MS; GC 8860 System-MSD 5977C, Agilent Technologies). The GC-MS utilized a DB-WAX Ultra Inert (30m, 0.25mm, 0.25µm capillary column, Agilent Technologies). 1μL of the sample was injected in split mode (10:1). Helium served as the carrier gas with a flow rate of 1.2 mL/min through the column. The injection, transfer line, quadrupole and ion source temperatures were 250 °C, 250 °C, 150 °C and 230 °C. The column temperature program started at 120 °C (held for 2 min), increased to 140 °C at a rate of 5 °C/min (held for 3 min), and then ramped to 250 °C at a rate of 20 °C/min, maintaining this temperature for 10 min. Electron impact mode was used with an energy of -70 eV. Mass spectrometry data were acquired in full scan mode within an *m/z* range of 35–550, with an acquisition frequency of 2.8 scans per second and a solvent delay of 2.8 min. Compound identification was confirmed by injecting pure standards and comparing retention times and corresponding MS spectra. Quantification of the analytes was performed in selected ion monitoring (SIM) mode using target ions for detection and confirmatory ions for validation (listed in Supplementary Table 1). Authentic FAs standards were prepared at concentrations ranging from 5 mM to 0.3125 mM (diluted 1:2 in water). Calibration curve aliquots underwent extraction alongside stool samples as described above, using 2-ethylbutyric acid as the internal standard. Data were processed in MassHunter software (Agilent Technologies), and blank values were subtracted from all measurements.

### LC-MS/MS analysis

The analytical system consists of a Dionex UltiMate 3000 HPLC (Thermo Fisher Scientific, Waltham, MA, USA) coupled to an AB Sciex 3200 QTRAP tandem mass spectrometer (Sciex, Framingham, MA, USA) operated in negative ESI mode. Mass spectrometry parameters are detailed in a prior study.^26^ Chromatographic separation was achieved on a Cortecs T3 column (2.1 × 100 mm, 2.7 µm; Waters, Milford, MA, USA) using mobile phases (A) water + 0.1% formic acid and (B) acetonitrile. The gradient program (%B) was 0–3 min 5%, 3–31 min 5–95%, 31–35 95% followed by 7 min of re-equilibration at 5%. The flow rate was 0.3 mL/min, the column and the autosampler temperature were 35 °C and 15 °C. Blank values were subtracted from the quantified concentrations in all samples.

### Statistical Analysis

All statistical analyses were performed in R (v4.4.1) using RStudio (Posit Team, 2025).^27^ Fatty acid concentrations were transformed on the natural log scale prior to modelling. To assess baseline differences across matrices or collection devices at t0, separate linear models were fitted for each fatty acid, with group (e.g. stool device or blood matrix) included as a fixed effect; pairwise contrasts between groups were then obtained using estimated marginal means (emmeans).^28^ At baseline, within-group associations between FA were explored by computing Spearman correlation matrices (Hmisc::*rcorr*).^29^ Correlation coefficients were visualised as clustered heatmaps to summarise the structure of co-variation in each matrix.

Longitudinal changes under RT storage were analysed using linear mixed-effects models fitted with *lmer* ^30^, with group, timepoint and their interaction as fixed effects and a random intercept for each subject (sample ID) to account for repeated measures. Post hoc comparisons between groups within timepoint and between timepoints within group were derived from the fitted models using the emmeans framework.^28^

Temporal trends were estimated by refitting mixed-effects models with time as a continuous variable. Because fatty acid concentrations were analysed on the natural log scale, exponentiation of the estimated time slopes yielded fold changes per unit of time, interpreted as multiplicative changes in concentration. Group-specific trends and 95% confidence intervals were extracted using the *emtrends* function. Estimated log-linear time slopes (β) for each fatty acid and sampling matrix, together with exponentiated fold changes per unit of time and 95% confidence intervals, are summarised in Supplementary Table S1.

Fatty acid profiles were evaluated using Bray-Curtis dissimilarities computed on log-transformed, total-sum-normalised data matrices. Profiles were visualised by principal coordinates analysis (PCoA), and differences between groups or timepoints were tested using permutational multivariate analysis of variance (PERMANOVA).^31^ For each timepoint, pairwise PERMANOVA was performed between groups, and for each group, pairwise comparisons were run between timepoints.^32^

Unless otherwise specified, all p-values were adjusted for multiple testing using the Benjamini–Hochberg false discovery rate (FDR), applied separately within each family of related hypotheses (e.g. baseline contrasts, time-trend estimates, and post hoc comparisons).

## RESULTS

### Faecal fatty acids baseline differences between collection devices

Principal coordinate analysis (PCoA) based on the faecal FA profiles at baseline (t0) revealed a clear separation according to the type of collection device (Fig. 2A). Samples collected with the OMNIgene-GUT device formed a distinct cluster compared with both untreated and eNAT-preserved faeces, which largely overlapped. PERMANOVA confirmed significant compositional differences between OMNIgene-GUT and the other two conditions (OMNIgene vs Untreated, p = 0.004; OMNIgene vs eNAT, p = 0.027), whereas no separation was detected between untreated and eNAT samples (p = 0.723). These results indicate that the overall faecal FA composition differs slightly depending on the preservation method used at the time of collection.

**Figure 2.**
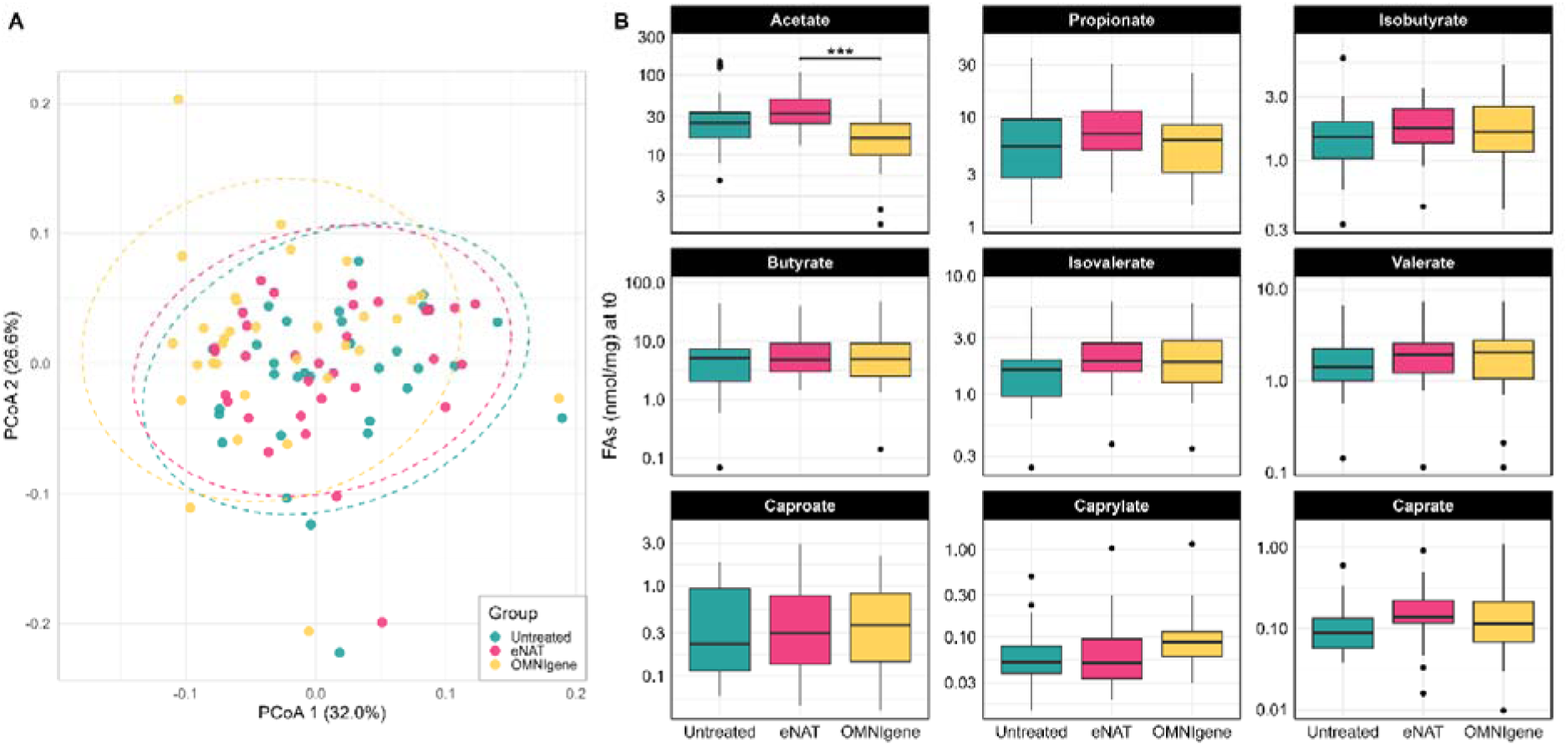
Baseline concentrations of faecal FAs across preservation conditions. **A**: Principal coordinates analysis (PCoA) based on Bray–Curtis dissimilarities of log-transformed FAs concentrations at baseline (t0). Samples cluster according to the collection device, with OMNIgene showing partial separation from untreated and eNAT. Ellipses represent 95% confidence intervals around group centroids. **B**: Concentrations (nmol/mg) of individual FAs at t0 across the three collection conditions. Among all metabolites, only acetate differed significantly between eNAT and OMNIgene (***p < 0.001). All other pairwise comparisons were not significant.

At the single-metabolite level, boxplot analysis (Fig. 2B) showed that the relative distribution of the nine measured FAs (acetate, propionate, isobutyrate, butyrate, isovalerate, valerate, caproate, caprylate, caprate) was broadly comparable across devices. Median concentrations were highest for acetate, propionate, and butyrate in all conditions, while branched- and medium-chain FAs displayed lower and more variable values. Among all compounds, only acetate showed a statistically significant difference across collection systems, with lower levels in OMNIgene-GUT compared with eNAT (p < 0.001) and a trend towards lower values relative to untreated stool (p = 0.054). No other metabolites exhibited significant pairwise differences between groups.

Correlation analyses at baseline revealed broadly similar patterns of co-variation among FAs across the three stool collection methods (Fig. 3). In untreated samples, the nine quantified FAs displayed a consistent positive correlation structure, with strong associations among the major SCFAs (acetate, propionate, butyrate) and moderate correlations extending to branched-chain FAs (BCFAs) and medium-chain species (MCFAs). A comparable pattern was observed in eNAT-preserved samples, where correlations remained uniformly positive and of similar magnitude, indicating that the stabilising buffer did not markedly alter the intrinsic relationships between metabolites. In OMNIgene samples, the overall structure was also dominated by positive associations, although several pairwise correlations appeared attenuated or shifted towards weaker values compared with untreated and eNAT, particularly for caprylate and caprate. Despite these minor differences, the global correlation profiles were largely conserved across devices.

**Figure 3.**
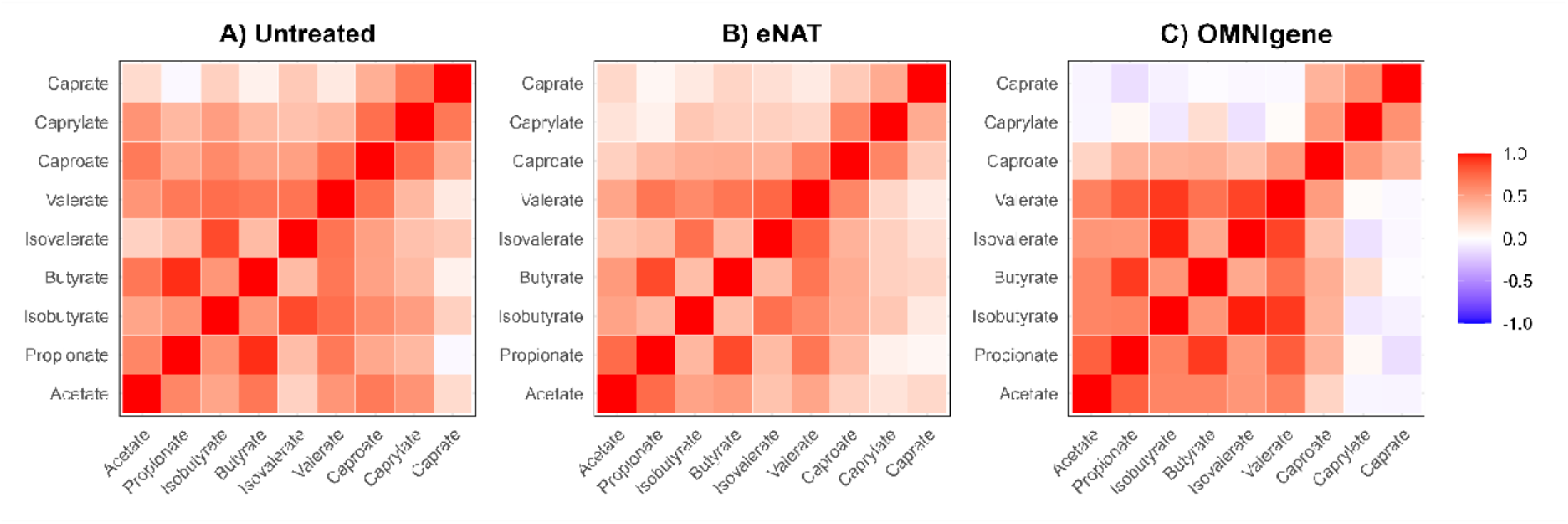
Spearman correlation matrices of FAs profiles at baseline (t0) across the three sampling methods: Untreated (**A**), eNAT (**B**), and OMNIgene (**C**). Each cell represents the pairwise correlation between FAs, ranging from −1 (blue) to +1 (red).

### Temporal variation and preservation efficiency in faecal sample

Temporal trends in faecal FAs concentrations were first assessed within each collection condition. In untreated faeces, the overall composition remained stable after freezing, as shown by the PCoA (Fig. 4A), where samples collected at baseline (t0) and after long-term storage at −80 °C (t4) largely overlapped without any detectable clustering by timepoint (p=0.507). Consistently, individual FA concentrations did not show marked deviations between t0 and t4 (Fig. 4B). Median values for acetate, propionate, and butyrate were comparable, while minor and medium-chain FAs displayed wider dispersion but no major shifts.

**Figure 4.**
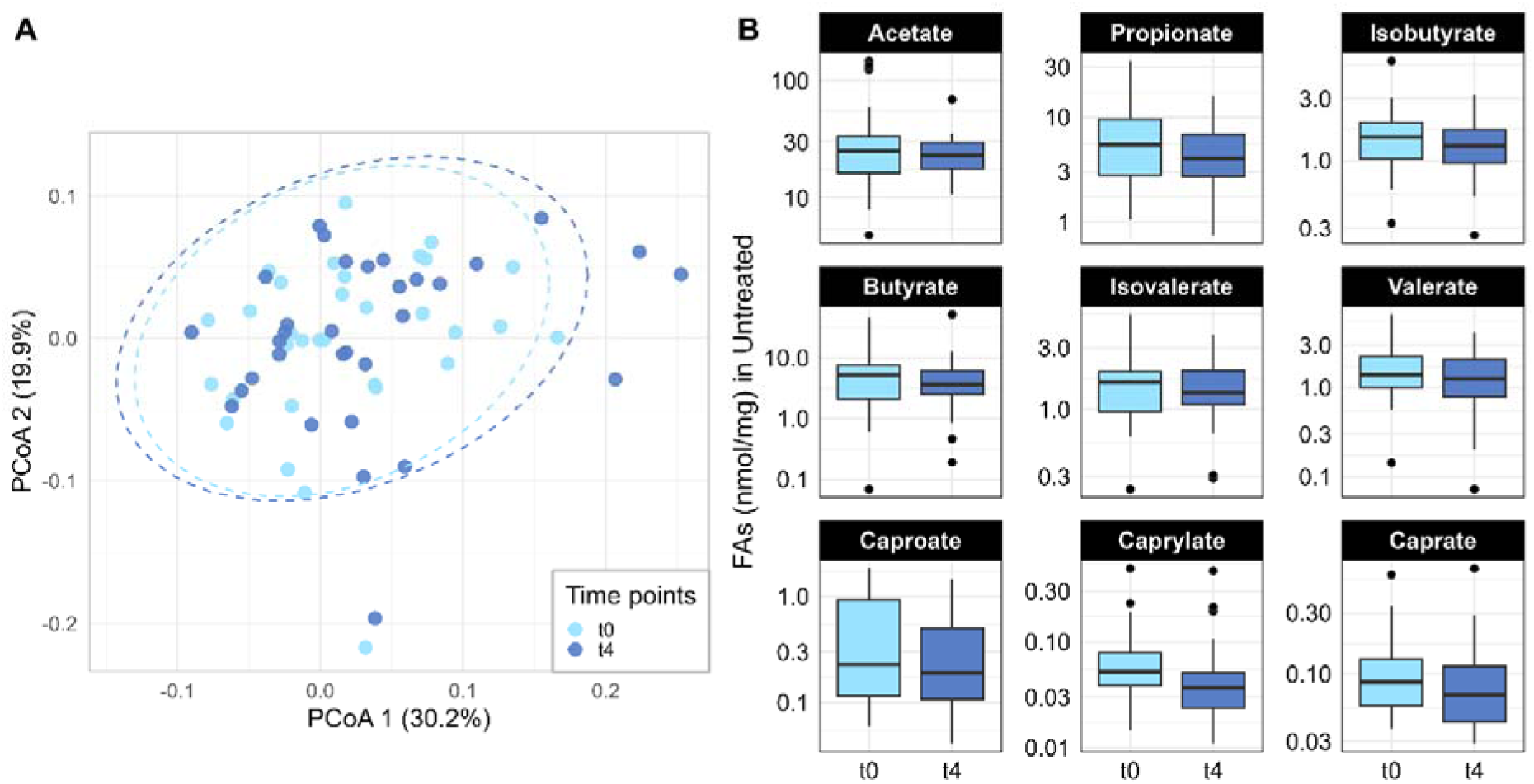
**A**: Principal coordinates analysis (PCoA) based on Bray–Curtis dissimilarities of FA profiles at baseline (t0) and after 21 days at −80 °C storage (t4). Samples from the two timepoints showed substantial overlap, with no evidence of major compositional shifts (p = 0.507). Ellipses indicate 95% confidence intervals. **B**: Concentrations (nmol/mg) of individual FAs in untreated stool at t0 and t4. None of comparisons were significant.

When the stability was evaluated under RT conditions, distinct trends emerged across preservation devices (Fig. 5). In untreated samples, all major SCFAs showed positive temporal slopes, indicating gradual increases during RT storage: acetate, propionate, isobutyrate, butyrate and isovalerate increased by approximately 5% per day (all p < 0.001), while valerate exhibited a more moderate but still significant rise (p = 0.007). In contrast, OMNIgene-stabilised samples exhibited generally stable profiles with limited variability across timepoints. Two compounds, however, deviated from this pattern: propionate (p = 0.010) and caprylate (p < 0.001) both showed negative slopes, reflecting a gradual decrease in concentration over time. For the eNAT device, most metabolites remained steady across the 21-day period, with only caprate (p < 0.001) showing a declining trend, while other FAs fluctuated within the expected technical range.

**Figure 5.**
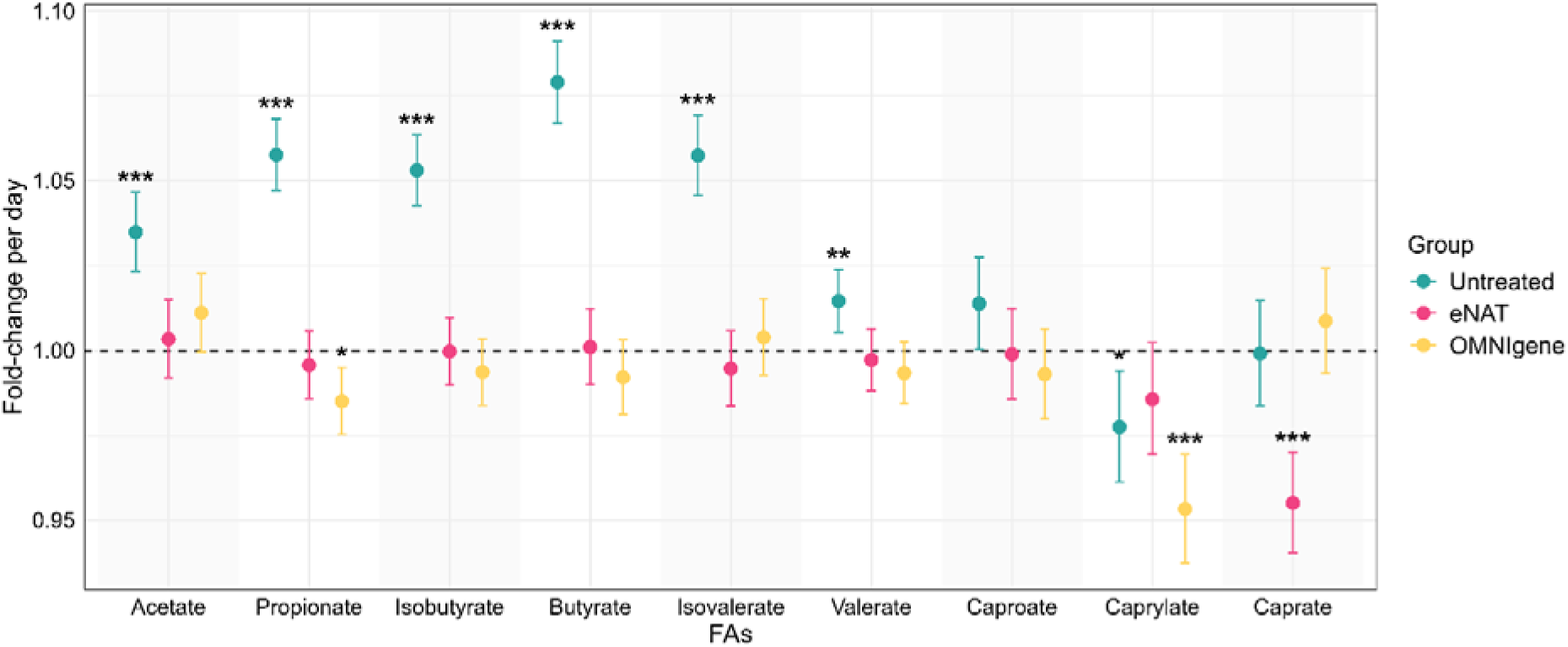
Temporal trends in faecal FAs across stool preservation methods. Fold-change per day derived from mixed-effects models for each FA in untreated, eNAT and OMNIgene samples stored at room temperature. Points represent estimated fold-change per day, and error bars indicate 95% confidence intervals. The dashed horizontal line denotes no change over time (fold-change = 1). Untreated samples show significant positive slopes for all major SCFAs, whereas eNAT samples remain close to unity for all analytes. In OMNIgene, most FAs are stable, with significant negative slopes for propionate, caprylate and caprate. Significance levels: *p < 0.05, **p < 0.01, ***p< 0.001.

### Systemic fatty acid baseline differences between matrices

To extend the analysis beyond the intestinal compartment, we next examined FA profiles in systemic matrices at baseline. Principal coordinates analysis revealed distinct clustering of plasma, whole blood and dried blood spots (DBS), indicating matrix-dependent compositional differences (Fig. 6A). Plasma samples formed a compact cluster and partially overlapped with whole blood (PERMANOVA p = 0.501). In contrast, DBS occupied a clearly separated region of ordination space and differed significantly from both blood (p = 0.002) and plasma (p = 0.002).

**Figure 6.**
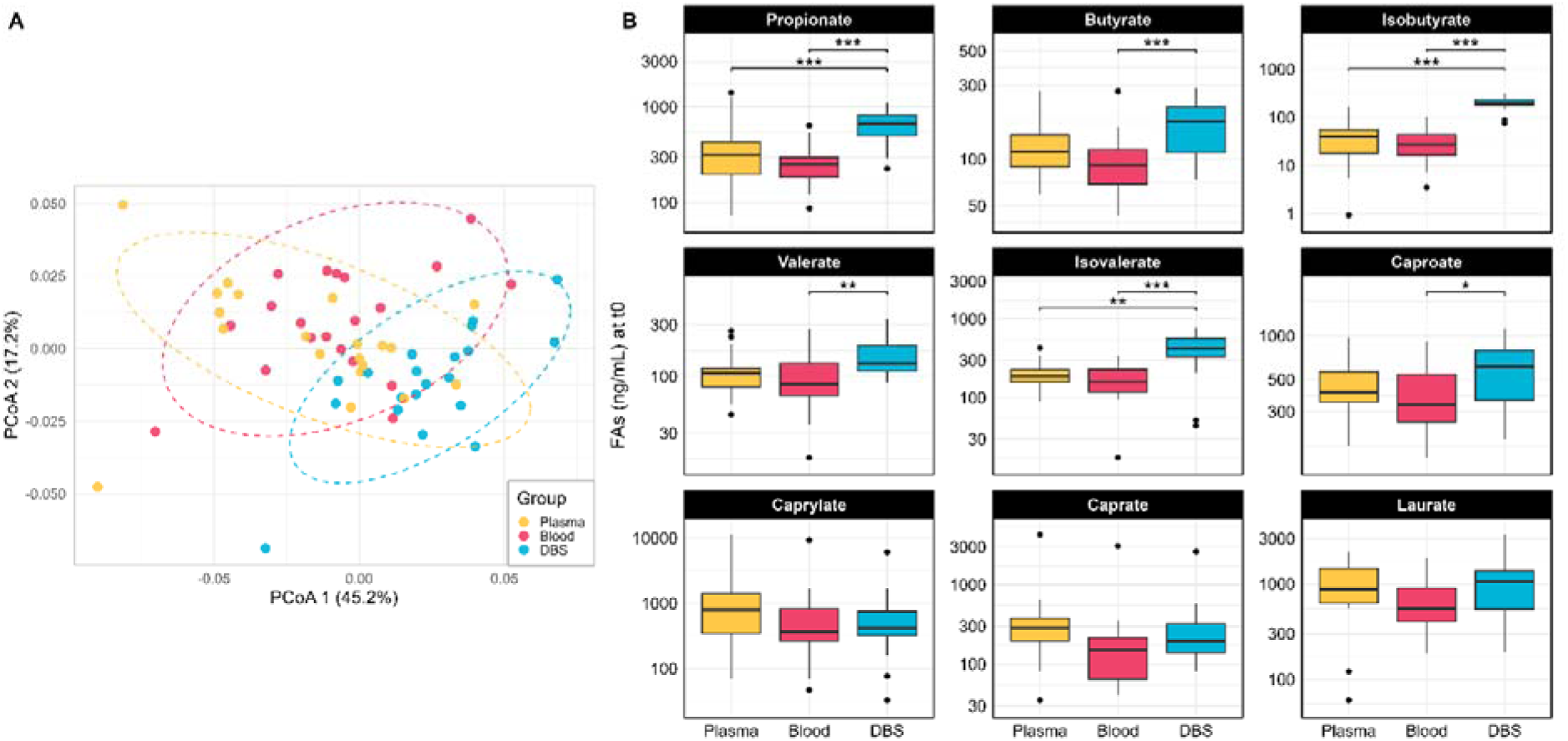
Baseline systemic fatty acid profiles across plasma, whole blood and DBS. **A**: Principal coordinates analysis (PCoA) of Bray–Curtis dissimilarities at baseline (T0). Plasma, whole blood and DBS form distinct clusters. Ellipses denote 95% confidence intervals. **B**: Boxplots showing the distribution of individual FAs across the three systemic matrices at T0. Several metabolites differed significantly between matrices, including propionate (Plasma vs DBS: p < 0.001 ; Blood vs DBS: p < 0.001), butyrate (Blood vs DBS: p < 0.001), isobutyrate (Plasma vs DBS: p < 0.001 ; Blood vs DBS: p < 0.001), valerate (p = 0.008), isovalerate (Plasma vs DBS: p = 0.008; Blood vs DBS: p < 0.001), and caproate (p = 0.034). Significance levels: *p < 0.05, **p < 0.01, ***p < 0.001.

Univariate comparisons further supported these matrix-specific signatures (Fig. 6B). DBS showed markedly higher concentrations of several SCFAs, including propionate (vs plasma: p < 0.001; vs blood: p < 0.001), butyrate (vs plasma: p = 0.091; vs blood: p < 0.001), isobutyrate (vs plasma: p < 0.001; vs blood: p < 0.001), valerate (vs plasma: p = 0.091; vs blood: p = 0.008), isovalerate (vs plasma: p = 0.008; vs blood: p < 0.001) and caproate (vs plasma: p = 0.224; vs blood: p = 0.034). Medium-chain fatty acids, including caprylate, caprate and laurate, did not differ substantially between whole blood and DBS, suggesting a more homogeneous distribution for these analytes across blood-derived matrices.

Correlation analyses at baseline revealed marked differences in the internal structure of FA co-variation across the three systemic matrices (Fig. 7). In plasma (Fig. 7A), SCFAs displayed a mixture of positive and negative associations and did not form a clearly defined cluster, resulting in a heterogeneous correlation pattern. By contrast, MCFAs exhibited a strong block of positive correlations, forming a coherent MCFA cluster that was not observed for the SCFAs. Whole blood (Fig. 7B) displayed a more homogeneous pattern, with uniformly positive correlations across nearly all analytes, suggesting a tighter inter-analyte coupling when cellular components are present. DBS samples (Fig. 7C) showed the strongest and most coherent correlation structure, characterised by consistently high positive correlations among SCFAs and several MCFAs, including laurate. Caprylate and caprate displayed weaker or slightly negative associations with SCFAs, but the overall matrix was dominated by strong positive co-variation.

**Figure 7.**
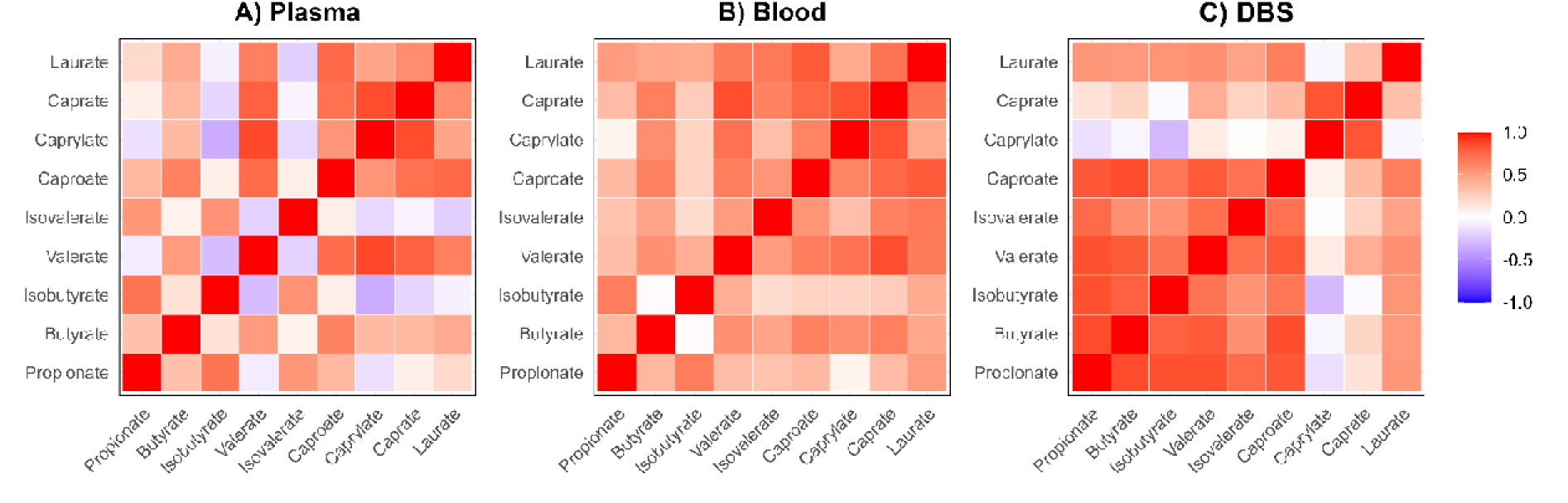
Baseline correlations among systemic FAs across plasma, whole blood and DBS. Correlation heatmaps showing Spearman coefficients between FAs in **(A)** plasma, **(B)** whole blood and **(C)** DBS at baseline. Colour scale represents Spearman’s ρ from −1 (blue) to +1 (red).

Temporal trends in systemic FAs were modelled over the RT interval from T0 to T14, revealing clear matrix-dependent patterns of stability (Fig. 8). In plasma, several analytes exhibited modest positive slopes consistent with gradual increases over time, including isobutyrate (p = 0.001), valerate (p < 0.001), caproate (p < 0.001) and caprylate (p = 0.008). Whole blood showed a distinct behaviour, characterised by uniformly positive temporal slopes across nearly all fatty acids. The magnitude of these increases, typically within the 5–15% per-day range, suggests a marked and systematic temporal drift in this matrix during the T0–T14 window. In contrast to this generally upward trend, caprate (p = 0.102) and laurate (p = 0.586) remained stable. DBS displayed bidirectional changes (Fig. 8): negative slopes for propionate, butyrate, isobutyrate, isovalerate, caprate, laurate; positive for valerate, caproate, caprylate. We next assessed long-term stability of FAs in DBS stored at RT up to 180 days (Fig. 9). Principal coordinates analysis revealed that DBS profiles remained largely stable from T14 to T180 (Fig. 9A, p = 0.129), with substantial overlap between the two timepoints and no evidence of major compositional drift. By contrast, samples collected at T0 formed a clearly separated cluster, indicating that RT storage introduces detectable shifts in FA composition (T0 vs T14: p = 0.001; T0 vs T180: p = 0.001). To quantify these trends, temporal slopes were modelled across the T14–T180 interval (Fig. 9B). Temporal slopes (fold-change/day) were near unity for most analytes, with only laurate showing a minor increase (p=0.002); all other SCFAs/MCFAs remained stable (p>0.05).

**Figure 8.**
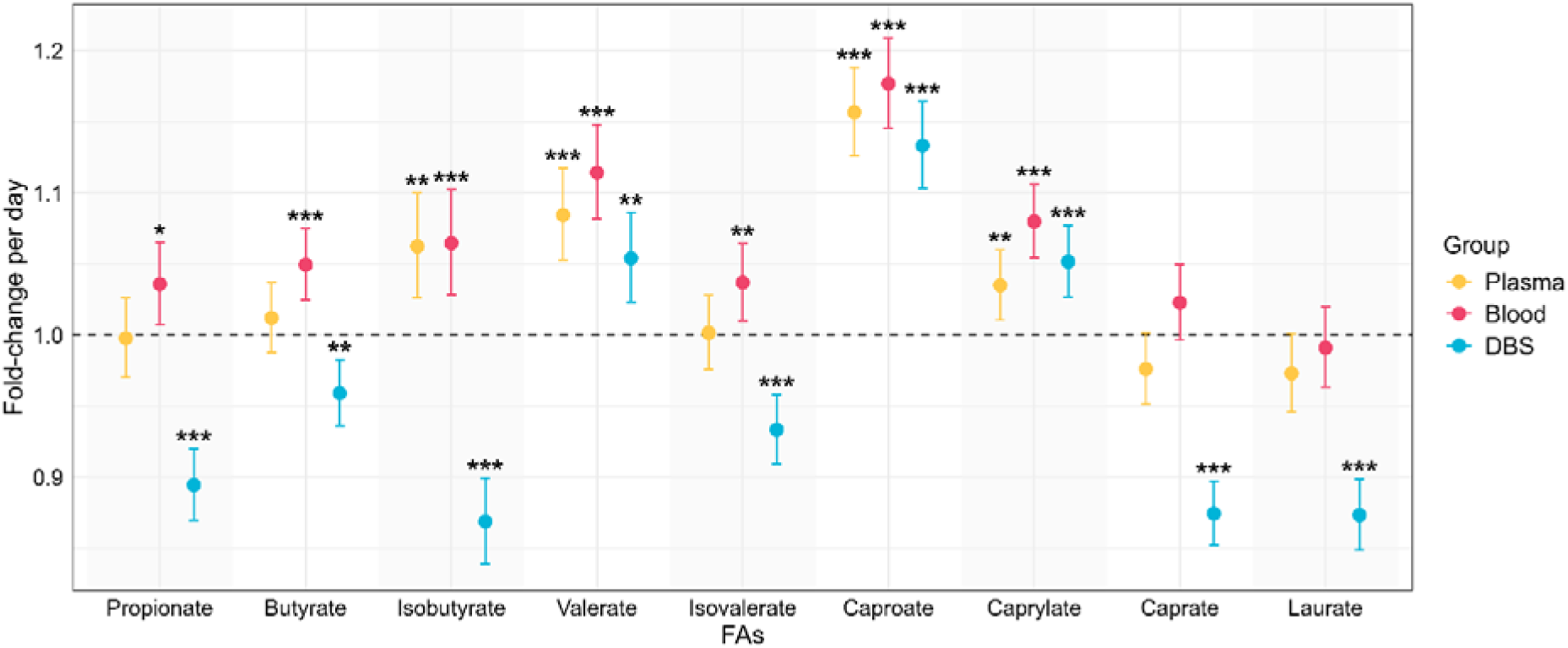
Temporal trends in systemic FAs across different matrices. Fold-change per day derived from mixed-effects models for each FA in plasma, whole blood and DBS samples stored at room temperature from T0 to T14. Points represent estimated fold-change per day, and error bars indicate 95% confidence intervals. The dashed horizontal line denotes no change over time (fold-change = 1). Significance levels: *p < 0.05, **p < 0.01, ***p< 0.001.

**Figure 9.**
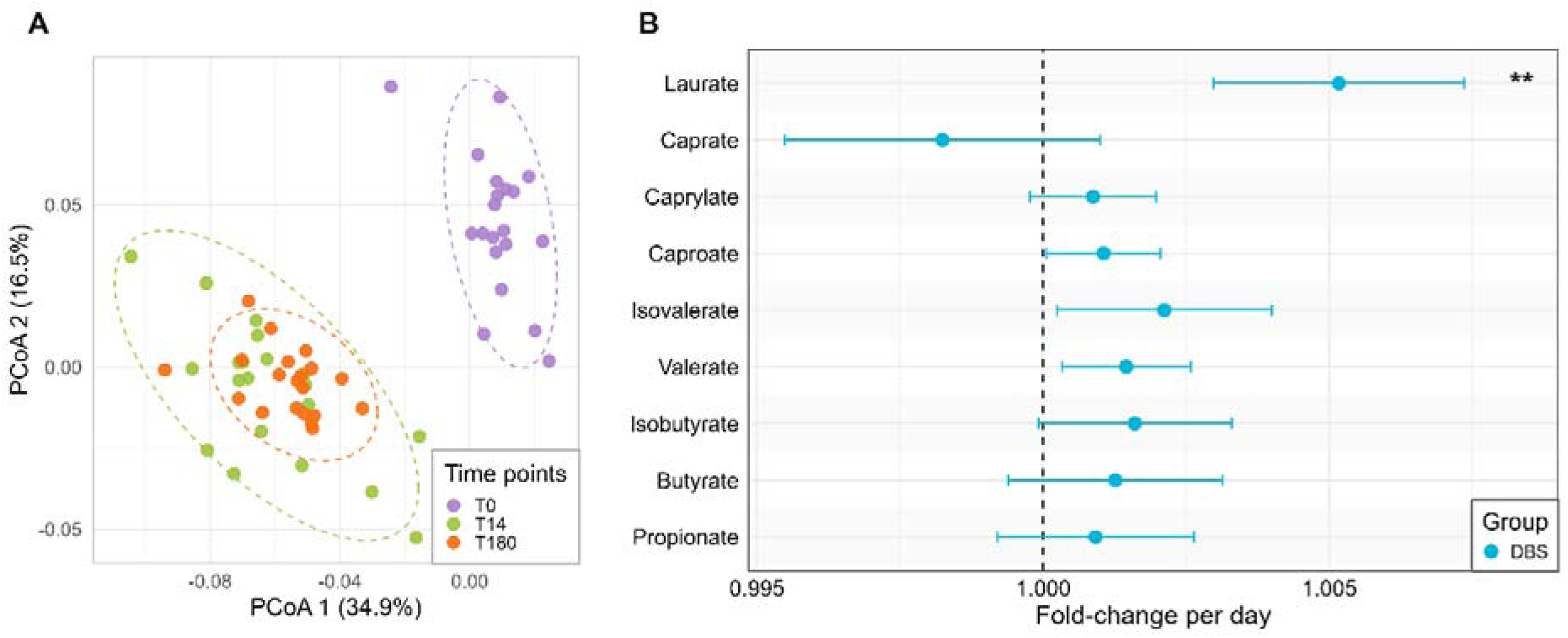
Long-term room-temperature stability of FAs in DBS. **A)** Principal coordinates analysis (Bray–Curtis dissimilarities) of DBS FA profiles at T0 (purple), T14 (green) and T180 (orange) shows a distinct separation at T0 and a substantial overlap between T14 and T180 (p = 0.129). **B)** Daily fold-changes estimated from mixed-effects models across the T14–T180 interval. Points represent fold-change per day, and error bars indicate 95% confidence intervals. Most analytes remained close to unity with only laurate showing a small but significant increase (p = 0.002).

## DISCUSSION

This study shows that microbiome study design, when encompassing microbial metabolites quantification is pivotal in guarantee accurate data. Indeed, sampling matrix and collection device have an important impact on the stability and interpretability of short- and medium-chain fatty acid profiles. By combining controlled RT storage experiments with complementary GC-MS and LC-MS/MS workflows, we provide a systematic assessment of how different non-invasive and user-friendly strategies preserve, distort or stabilise FA signatures over time. These findings have direct implications for microbiome-metabolome research, where SCFAs are increasingly used as functional signature of microbial activity. Moreover, the self-collection and the easy (RT) shipment may reduce the drop out phenomenon in longitudinal and multicentric studies.

For faecal samples, our data reveal marked differences between the investigated stabilisation devices in their ability to preserve lipid profiles. Both eNAT and OMNIgene-GUT were originally developed for the preservation of nucleic acids and are widely used in microbiome research to stabilise microbial DNA during transport and storage.^20,33–35^ While their performance for genomic applications has been documented, their suitability for preserving volatile metabolites has not been systematically evaluated.

In addition to nucleic acid preservation, eNAT has also been used in metaproteomic workflows, suggesting a broad range of omics applications.^36^ OMNIgene-GUT has likewise been explored in selected studies assessing SCFA and metabolite quantification; however, evidence remains limited and largely restricted to a subset of major metabolites.^37–39^ Notably, the existence of dedicated metabolite-preserving collection systems from the same manufacturer (e.g., OMNImet®•GUT, DNA Genotek) suggests that nucleic acid stabilisation buffers may not fully address the specific requirements of metabolomic analyses.^40^ This consideration is particularly relevant for volatile compounds such as SCFAs, whose chemical stability differs fundamentally from that of DNA.

To our knowledge, this is the first study to evaluate eNAT as a collection device for FA analysis. In this context, eNAT proved highly effective: both multivariate structure and individual FA concentrations closely matched the untreated baseline (t0), and temporal stability at RT was maintained for up to 21 days, except for caprate. A plausible explanation lies in the chemical composition of eNAT, which contains guanidine thiocyanate and other denaturing agents capable of rapidly lysing microbial cells and inhibiting enzymatic activity.^33^ By effectively suppressing post-collection microbial metabolism, eNAT appears to preserve the in vivo SCFA profile even under prolonged ambient storage. These results position eNAT as a robust option for faecal FA analysis, particularly in settings where immediate freezing is impractical.

In contrast, OMNIgene-GUT displayed a distinct and more complex behaviour. At baseline, its lipid profile diverged from both untreated stool and eNAT-treated samples, indicating that the buffer alters the measurable FA pool before storage begins. Consistent with a previous report,^39^ the concentrations of the three major SCFAs were comparable to untreated stool; however, broader FA profiling revealed pronounced baseline differences, suggesting that OMNIgene-GUT may influence extraction efficiency or chemical recoverability for less commonly measured species. Over time, OMNIgene-GUT also exhibited reduced stability, with significant declines in propionate and caprylate during RT storage.

These observations suggest that, although OMNIgene-GUT effectively preserves genetic material, it does not fully prevent chemical or microbial processes affecting FA profiles.^34,37,41^ McKay et al.^42^ reported stable SCFA measurements from OMNIgene-GUT samples following chemical derivatisation, a well-established strategy for stabilising volatile metabolites. Importantly, this suggests that the apparent robustness of SCFA profiles observed in this study may partially reflect the stabilising effect of derivatisation rather than intrinsic preservation by the collection device alone. Taken together, our results indicate that OMNIgene-GUT introduces both baseline and temporal alteration in FA measurements, whereas eNAT provides more consistent preservation of faecal lipid profiles under ambient conditions.

As expected, untreated stool stored at RT exhibited substantial increases in major SCFAs over time, consistent with the well-documented continuation of microbial fermentation ex vivo. Faecal samples remain metabolically active outside the gut environment, with measurable changes occurring within hours, even at moderate temperatures.^8,16,43^ Our findings are consistent with the established observation that all major SCFAs increased over time, until microbial activity was presumably limited by substrate depletion or processing.^10,44,45^ Importantly, immediate freezing at −80 °C effectively preserved the SCFA profile, as no significant differences were observed between fresh t0 samples and frozen aliquots re-analysed at the end of the study. This confirms that rapid freezing remains a gold standard for minimising fermentation-driven artefacts in stool metabolomics.

In systemic compartments, this study provides a structured evaluation of DBS for SCFA and MCFA analysis over an extended RT storage interval, an area that has been minimally explored, as DBS have been used in multi-matrix SCFA workflows without systematic assessment of their temporal stability.^46,47^ DBS sampling requires minimal blood volume, is minimally invasive, allow self-sampling, and is well suited to decentralised or longitudinal study designs, making it an attractive alternative to conventional venous sampling.^21,48,49^

At baseline, DBS differed from plasma and whole blood, as reflected by clear separation in PCoA space and by higher concentrations of several SCFAs. These differences may arise from a combination of matrix-specific extraction efficiencies, concentration effects during drying, and altered protein-binding dynamics. Notably, despite differences in absolute abundance, the correlation structure of DBS closely resembled that of whole blood, indicating that relative relationships among FA species are largely preserved. This suggests that DBS can capture a coherent metabolic fingerprint, even when absolute concentrations differ from liquid matrices. Future calibration studies integrating controlled spike-in designs and matrix-specific recovery profiling may elucidate whether robust and biologically meaningful conversion factors can be established between DBS and whole blood.

RT stability in systemic matrices proved strongly matrix-dependent. Both plasma and whole blood exhibited consistent upward drift over time, with multiple SCFAs and MCFAs increasing during storage. This behaviour has been reported previously and may reflect ongoing enzymatic activity, erythrocyte-derived release, or chemical conversion of precursor molecules.^50^ Erythrocytes are particularly sensitive to temperature-induced stress, and haemolysis can release intracellular enzymes and iron, both of which can promote lipid peroxidation cascades.^51–53^ One possible consequence is the oxidative degradation of long-chain polyunsaturated fatty acids, potentially generating shorter-chain oxidation products,^54,55^ although this mechanism remains speculative and warrants further investigation.

In contrast, most analytes in DBS declined over time, likely due to a combination of oxidative loss, evaporation and interactions with the cellulose matrix.^46,56^ Unlike liquid matrices, DBS showed an opposite trajectory, stabilizing after ∼14 days RT storage. When stored for up to six months, DBS samples clustered tightly with their day-14 counterparts in multivariate space, and only laurate continued to show significant drift. This apparent plateau suggests that DBS reach a physicochemical equilibrium state, after which further degradation becomes negligible. Chemical derivatisation is commonly used to improve the stability of volatile metabolites such as SCFAs, but it adds complexity to sample preparation and is typically justified when degradation is ongoing.^57–59^ In the present study, the post-day-14 stability observed in DBS indicates that derivatisation may not be necessary for long-term ambient storage once the dried matrix has equilibrated. However, the initial decline observed during the first 14 days suggests that early storage conditions may influence metabolite stability. It is conceivable that refrigeration at 4 °C or immediate freezing at −80 °C during this initial phase could mitigate early degradation, although this was not directly assessed in the present study. Once equilibrium is reached, DBS appear to provide sufficient protection to preserve FA profiles for extended periods, offering a practical and low-maintenance solution for large-scale metabolomic studies.

## CONCLUSION

Together, these findings emphasise that fatty acid measurements are highly sensitive to pre-analytical decisions and that these choices have lasting consequences for data interpretation. Importantly, once a sampling strategy is selected, it defines the trajectory of the resulting fatty acid profile: samples collected under different conditions do not converge over time and should therefore not be treated as interchangeable.

Our results further demonstrate that adherence to defined storage timeframes is critical. Deviations from recommended time windows introduce systematic, matrix-dependent changes that cannot be fully corrected at later stages of analysis. Respecting storage duration and conditions is therefore essential to preserve biologically meaningful signals, particularly when fatty acids are used as functional readouts of microbial activity.

Finally, the choice of collection device itself constitutes a biological decision rather than a neutral technical detail. Different devices variably stabilise or distort fatty acid profiles, shaping both baseline measurements and temporal dynamics. Awareness of these effects is crucial when designing studies, comparing datasets, or interpreting microbiome-metabolome associations.

Taken together, this work provides practical guidance for the optimal use of faecal and systemic sampling tools in microbiome-associated fatty acid studies. By aligning sampling strategy, storage conditions and analytical interpretation, future studies can improve reproducibility and ensure that observed differences reflect biological variation rather than pre-analytical artefacts.

## Funding

This work was supported by the PNRR “D34Health - Digital Driven Diagnostics, prognostics and therapeutics for sustainable Health care” Grant (# PNC0000001).

## Disclosure statement

The authors report there are no competing interests to declare.

## Data availability statement

The authors confirm that the data supporting the findings of this study are available within the article and its supplementary materials. The datasets generated and analysed during the current study will be deposited in the Zenodo repository and made publicly available upon publication (DOI: 10.5281/zenodo.18864940).

## Notes

### Competing Interest Statement

The authors have declared no competing interest.

### Author Declarations

The study was reviewed and approved by the Comitato Etico Territoriale Lombardia 1 (approval number CET 129-2024). All participants provided written informed consent, and the study was conducted in accordance with the Declaration of Helsinki.

## REFERENCES

1. Silva YP, Bernardi A, Frozza RL. The Role of Short-Chain Fatty Acids From Gut Microbiota in Gut-Brain Communication. Front Endocrinol (Lausanne). Frontiers Media S.A. 2020;11. doi:10.3389/fendo.2020.00025

2. Dalile B, Van Oudenhove L, Vervliet B, Verbeke K. The role of short-chain fatty acids in microbiota–gut–brain communication. Nat Rev Gastroenterol Hepatol. Nature Publishing Group. 2019;16(8):461–478. doi:10.1038/s41575-019-0157-3

3. Koh A, De Vadder F, Kovatcheva-Datchary P, Bäckhed F. From Dietary Fiber to Host Physiology: Short-Chain Fatty Acids as Key Bacterial Metabolites. Cell. 2016;165(6):1332–1345. doi:10.1016/j.cell.2016.05.041

4. Blaak EE, Canfora EE, Theis S, et al. Short chain fatty acids in human gut and metabolic health. Benef Microbes. Wageningen Academic Publishers. 2020;11(5):411–455. doi:10.3920/BM2020.0057

5. Visconti A, Le Roy CI, Rosa F, et al. Interplay between the human gut microbiome and host metabolism. Nat Commun. 2019;10(1). doi:10.1038/s41467-019-12476-z

6. Tremaroli V, Bäckhed F. Functional interactions between the gut microbiota and host metabolism. Nature. 2012;489(7415):242–249. doi:10.1038/nature11552

7. Mayo-Martínez L, Paz Lorenzo M, Martos-Moreno G, et al. Short-chain fatty acids in plasma and feces: An optimized and validated LC-QqQ-MS method applied to study anorexia nervosa. Microchemical Journal. 2024;200(9):110255. doi:10.1016/j.microc.2024.110255

8. Cunningha JL, Bramstång L, Singh A, et al. Impact of time and temperature on gut microbiota and SCFA composition in stool samples. PLoS One. 2020;15(8 August). doi:10.1371/journal.pone.0236944

9. Vandeputte D, Falony G, Vieira-Silva S, Tito RY, Joossens M, Raes J. Stool consistency is strongly associated with gut microbiota richness and composition, enterotypes and bacterial growth rates. Gut. 2016;65(1):57–62. doi:10.1136/gutjnl-2015-309618

10. Liebisch G, Ecker J, Roth S, et al. Quantification of fecal short chain fatty acids by liquid chromatography tandem mass spectrometry—investigation of pre-analytic stability. Biomolecules. 2019;9(4). doi:10.3390/biom9040121

11. Weng CYC, Suarez C, Cheang SE, et al. Quantifying Gut Microbial Short-Chain Fatty Acids and Their Isotopomers in Mechanistic Studies Using a Rapid, Readily Expandable LC-MS Platform. Anal Chem. 2024;96(6):2415–2424. doi:10.1021/acs.analchem.3c04352

12. Fristedt R, Ruppert V, Trower T, Cooney J, Landberg R. Quantitation of circulating short-chain fatty acids in small volume blood samples from animals and humans. Talanta. 2024;272(6086):125743. doi:10.1016/j.talanta.2024.125743

13. Chalova P, Tazky A, Skultety L, et al. Determination of short-chain fatty acids as putative biomarkers of cancer diseases by modern analytical strategies and tools: a review. Front Oncol. Frontiers Media SA. 2023;13. doi:10.3389/fonc.2023.1110235

14. Mahdi T, Desmons A, Krasniqi P, et al. Effect of Stool Sampling on a Routine Clinical Method for the Quantification of Six Short Chain Fatty Acids in Stool Using Gas Chromatography–Mass Spectrometry. Microorganisms. 2024;12(4). doi:10.3390/microorganisms12040828

15. Anton G, Wilson R, Yu ZH, et al. Pre-analytical sample quality: Metabolite ratios as an intrinsic marker for prolonged room temperature exposure of serum samples. PLoS One. 2015;10(3). doi:10.1371/journal.pone.0121495

16. Mahdi T, Desmons A, Krasniqi P, et al. Effect of Stool Sampling on a Routine Clinical Method for the Quantification of Six Short Chain Fatty Acids in Stool Using Gas Chromatography–Mass Spectrometry. Microorganisms. 2024;12(4). doi:10.3390/microorganisms12040828

17. Ueyama J, Oda M, Hirayama M, et al. Freeze-drying enables homogeneous and stable sample preparation for determination of fecal short-chain fatty acids. Anal Biochem. 2020;589:113508. doi:10.1016/j.ab.2019.113508

18. Vandeputte D, Tito RY, Vanleeuwen R, Falony G, Raes J. Practical considerations for large-scale gut microbiome studies. FEMS Microbiol Rev. Oxford University Press. 2017;41:S154–S167. doi:10.1093/FEMSRE/FUX027

19. Gupta K, Mahajan R. Applications and diagnostic potential of dried blood spots. Int J Appl Basic Med Res. 2018;8(1):1. doi:10.4103/ijabmr.ijabmr_7_18

20. Richard-Greenblatt M, Comar CE, Flevaud L, et al. Copan eNAT transport system to address challenges in COVID-19 diagnostics in regions with limited testing access. J Clin Microbiol. 2021;59(5). doi:10.1128/JCM.00110-21

21. Triva F, Borghi E, Marsiglia MD, et al. Targeting the gut to improve seizure control in CDKL5 deficiency disorder (CDD): study protocol for a single-arm, open-label clinical trial. Front Neurol. 2025;16. doi:10.3389/fneur.2025.1642329

22. Abrahamson M, Hooker E, Ajami NJ, Petrosino JF, Orwoll ES. Successful collection of stool samples for microbiome analyses from a large community-based population of elderly men. Contemp Clin Trials Commun. 2017;7(5):158–162. doi:10.1016/j.conctc.2017.07.002

23. Tosi M, Marsiglia MD, Ottaviano E, et al. Gut Microbiota and Metabolic Modulation by Slow-Release Protein Substitutes in Phenylketonuria: Findings from the PREMP Study. Nutrients . 2025;17(24). doi:10.3390/nu17243829

24. Moat SJ, Dibden C, Tetlow L, et al. Effect of blood volume on analytical bias in dried blood spots prepared for newborn screening external quality assurance. Bioanalysis. 2020;12(2):99–109. doi:10.4155/bio-2019-0201

25. Hewawasam E, Liu G, Jeffery DW, Gibson RA, Muhlhausler BS. Estimation of the Volume of Blood in a Small Disc Punched From a Dried Blood Spot Card. European Journal of Lipid Science and Technology. 2018;120(3). doi:10.1002/ejlt.201700362

26. Dei Cas M, Paroni R, Saccardo A, et al. A straightforward LC-MS/MS analysis to study serum profile of short and medium chain fatty acids. Journal of Chromatography B. 2020;1154:121982. doi:10.1016/j.jchromb.2020.121982

27. R Core Team. R: A Language and Environment for Statistical Computing. https://www.R-project.org/. Preprint posted online 2024:4.4.1. Accessed February 18, 2026. https://www.R-project.org/

28. Lenth R V., Piaskowski J, Banfai B, et al. emmeans: Estimated Marginal Means, aka Least-Squares Means. Preprint posted online 2025. doi:10.32614/CRAN.package.emmeans

29. Harrell FE, Beck C, Dupont C. Hmisc: Harrell Miscellaneous. Preprint posted online 2026:5.2-5. doi:10.32614/CRAN.package.Hmisc

30. Bates D, Mächler M, Bolker BM, Walker SC. Fitting linear mixed-effects models using lme4. J Stat Softw. 2015;67(1). doi:10.18637/jss.v067.i01

31. Oksanen J, Simpson GL, Blanchet G, et al. vegan: Community Ecology Package. Preprint posted online 2025:2.7–2. doi:10.32614/CRAN.package.vegan

32. Martinez Arbizu P. pairwiseAdonis: Pairwise multilevel comparison using adonis. Preprint posted online 2020:0.4. Accessed February 18, 2026. https://github.com/pmartinezarbizu/pairwiseAdonis

33. Young RR, Jenkins K, Araujo-Perez F, Seed PC, Kelly MS. Long-term stability of microbiome diversity and composition in fecal samples stored in eNAT medium. Microbiologyopen. 2020;9(7). doi:10.1002/mbo3.1046

34. Lim MY, Hong S, Kim BM, Ahn Y, Kim HJ, Nam Y Do. Changes in microbiome and metabolomic profiles of fecal samples stored with stabilizing solution at room temperature: a pilot study. Sci Rep. 2020;10(1). doi:10.1038/s41598-020-58719-8

35. Hoogendijk R, van den Broek TJM, Lee H, et al. Omnigene-Guttm ensures fecal microbiome stability in the pediatric population. AMB Express. 2024;14(1). doi:10.1186/s13568-024-01798-x

36. Tanca A, Schallert K, Grenga L, et al. Critical Assessment of MetaProteome Investigation 2 (CAMPI-2): multi-laboratory assessment of sample processing methods to stabilize fecal microbiome for functional analysis. Microbiome. 2025;13(1). doi:10.1186/s40168-025-02248-x

37. Guan H, Pu Y, Liu C, et al. Comparison of Fecal Collection Methods on Variation in Gut Metagenomics and Untargeted Metabolomics. mSphere. 2021;6(5). doi:10.1128/msphere.00636-21

38. Takagi T, Kunihiro T, Takahashi S, et al. A newly developed solution for the preservation of short-chain fatty acids, bile acids, and microbiota in fecal specimens. J Clin Biochem Nutr. 2023;72(3):263–269. doi:10.3164/JCBN.22-107

39. Wang Z, Zolnik CP, Qiu Y, et al. Comparison of fecal collection methods for microbiome and metabolomics studies. Front Cell Infect Microbiol. 2018;8(AUG). doi:10.3389/fcimb.2018.00301

40. Cleminson J, Kerbiriou C, McKirdy S, et al. OMNImet®•GUT collection kit supports sample collection and ambient temperature preservation of faecal short chain fatty acids. Metabolomics. Springer. 2025;21(5). doi:10.1007/s11306-025-02322-3

41. Gemmell MR, Jayawardana T, Koentgen S, et al. Optimised human stool sample collection for multi-omic microbiota analysis. Sci Rep. 2024;14(1). doi:10.1038/s41598-024-67499-4

42. McKay MJ, Castaneda M, Catania S, et al. Quantification of short-chain fatty acids in human stool samples by LC-MS/MS following derivatization with aniline analogues. Journal of Chromatography B. 2023;1217:123618. doi:10.1016/j.jchromb.2023.123618

43. Isokääntä H, Pinto da Silva L, Karu N, et al. Comparative Metabolomics and Microbiome Analysis of Ethanol versus OMNImet/gene•GUT Fecal Stabilization. Anal Chem. 2024;96(22):8893–8904. doi:10.1021/acs.analchem.3c04436

44. Czarnowski P, Mikula M, Ostrowski J, Żeber-Lubecka N. Gas Chromatography–Mass Spectrometry-Based Analyses of Fecal Short-Chain Fatty Acids (SCFAs): A Summary Review and Own Experience. Biomedicines. Multidisciplinary Digital Publishing Institute (MDPI). 2024;12(8). doi:10.3390/biomedicines12081904

45. Lewandowska I, Grzech K, Krzysztoń-Russjan J. Importance of Human Faecal Biobanking: From Collection to Storage. Advancements of Microbiology. 2024;63(4):181–189. doi:10.2478/am-2024-0015

46. Palmer EA, Cooper HJ, Dunn WB. Investigation of the 12-Month Stability of Dried Blood and Urine Spots Applying Untargeted UHPLC-MS Metabolomic Assays. Anal Chem. Published online 2019. doi:10.1021/acs.analchem.9b02577

47. Monteiro J, Lefèvre A, Dufour-Rainfray D, et al. Multi-Compartment SCFA Quantification in Human. Am J Analyt Chem. 2024;15(06):177–200. doi:10.4236/ajac.2024.156012

48. Hewawasam E, Liu G, Jeffery DW, Gibson RA, Muhlhausler BS. Estimation of the Volume of Blood in a Small Disc Punched From a Dried Blood Spot Card. European Journal of Lipid Science and Technology. 2018;120(3). doi:10.1002/ejlt.201700362

49. Bossi E, Limo E, Pagani L, et al. Revolutionizing Blood Collection: Innovations, Applications, and the Potential of Microsampling Technologies for Monitoring Metabolites and Lipids. Metabolites. Multidisciplinary Digital Publishing Institute (MDPI). 2024;14(1). doi:10.3390/metabo14010046

50. Metherel AH, Stark KD. The stability of blood fatty acids during storage and potential mechanisms of degradation: A review. Prostaglandins Leukot Essent Fatty Acids. 2016;104:33–43. doi:10.1016/j.plefa.2015.12.003

51. Gordon Bell J, MacKinlay EE, Dick JR, Younger I, Lands B, Gilhooly T. Using a fingertip whole blood sample for rapid fatty acid measurement: Method validation and correlation with erythrocyte polar lipid compositions in UK subjects. British Journal of Nutrition. 2011;106(9):1408–1415. doi:10.1017/S0007114511001978

52. Lin YH, Hanson JA, Strandjord SE, et al. Fast transmethylation of total lipids in dried blood by microwave irradiation and its application to a population study. Lipids. 2014;49(8):839–851. doi:10.1007/s11745-014-3918-3

53. Kristensen NB. Quantification of whole blood Short-chain fatty acids by gas chromatographic determination of plasma 2-chloroethyl derivatives and correction for dilution space in erythrocytes. Acta Agriculturae Scandinavica A: Animal Sciences. 2000;50(4):231–236. doi:10.1080/090647000750069421

54. Guichardant M, Chen P, Liu M, et al. Functional lipidomics of oxidized products from polyunsaturated fatty acids. Chem Phys Lipids. 2011;164(6):544–548. doi:10.1016/j.chemphyslip.2011.05.002

55. Passi S, Picardo M, De Luca C, Nazzaro-Porro M, Rossi L, Rotilio G. Saturated dicarboxylic acids as products of unsaturated fatty acid oxidation. Biochimica et Biophysica Acta (BBA) - Lipids and Lipid Metabolism. 1993;1168(2):190–198. doi:10.1016/0005-2760(93)90124-R

56. Cui HN, Shi F, Huang G, et al. Evaluation of metabolite stability in dried blood spot stored at different temperatures and times. Sci Rep. 2024;14(1). doi:10.1038/s41598-024-82041-2

57. Fristedt R, Ruppert V, Trower T, Cooney J, Landberg R. Quantitation of circulating short-chain fatty acids in small volume blood samples from animals and humans. Talanta. 2024;272:125743. doi:10.1016/j.talanta.2024.125743

58. Trivedi N, Erickson HE, Bala V, Chhonker YS, Murry DJ. A Concise Review of Liquid Chromatography-Mass Spectrometry-Based Quantification Methods for Short Chain Fatty Acids as Endogenous Biomarkers. Int J Mol Sci. MDPI. 2022;23(21). doi:10.3390/ijms232113486

59. Xiang L, Zhu L, Huang Y, Cai Z. Application of Derivatization in Fatty Acids and Fatty Acyls Detection: Mass Spectrometry-Based Targeted Lipidomics. Small Methods. John Wiley and Sons Inc. 2020;4(8). doi:10.1002/smtd.202000160

